# Laromestrocel preserves brain volume and cognitive function in Alzheimer’s disease by inhibiting neuroinflammation

**DOI:** 10.64898/2026.09.16.26363132

**Authors:** Brian G. Rash, Sofya Levitina, Jeffrey Botbyl, Chris Conklin, Madhura Ingalhalikar, Joshua M. Hare

## Abstract

Neuroinflammation is a principal driver of brain atrophy and cognitive decline in Alzheimer’s disease (AD). Here we show that laromestrocel, a mesenchymal stem cell therapy, inhibits progressive brain inflammation in subjects (n=49) with mild AD, with the strongest effect seen in core AD brain regions: the left hippocampus (25 million cells (M)x4 monthly dose group; p<0.001, N=11, and 100Mx4 group; p<0.001; N=10), the left temporal cortex (25Mx4 group; p=0.007; N=11) and left parietal cortex (25Mx4, week 26; p=0.042; N=10), and the medulla (25Mx4 group and 100Mx4; p=0.007; N=11 and N=10) after 39 weeks compared with placebo (N=9). Reduced neuroinflammation correlated with reduced brain atrophy (hippocampus: R=−0.326; p=0.040; N=40) and improvements in clinical scores, particularly cognitive function (hippocampus x MoCA: R=−0.401; p=0.011; N=40). Treated subjects also exhibited reduced neurogranin (25Mx4 group; p=0.023; N=11) in blood plasma, consistent with reduced synaptic loss. Finally, blood biomarker analysis supported a convergent multimodal mechanism of action implicating peripheral immune engagement. Together these results show that long-lasting anti-neuroinflammatory effects of laromestrocel mitigate AD brain inflammation and are associated with markedly reduced brain tissue destruction and improved clinical function.

## Introduction

Neuroinflammation is a core neuropathological driver of Alzheimer’s disease (AD) ^1–5^. While accumulating amyloid and tau fibrils may trigger the disease, a key ensuing mechanism underlying AD brain damage is a hyperinflammatory state affecting synapses, neurites, and neuronal survival^6–8^. Various clinical and preclinical approaches have attempted to target this core AD driver, including microglial modulation via TREM2 activation^9^, cytokine targeting employing TNFα blockade^10^, complement system inhibition (preclinical stage development^11^), metabolic-immune modulation via GLP-1 receptor agonists^12,13^, and general NSAID administration^14^, among others. So far, none of these approaches has yielded a promising risk-efficacy profile. Only anti-amyloid immunotherapies have provided some therapeutic benefit to AD patients and their families by offering up to a 30% slowing of AD disease progression^15–18^; yet this benefit does not slow brain tissue destruction and is associated with a risk of amyloid-related imaging abnormalities (ARIA), which involve either brain swelling or bleeding that can be fatal^19–23^. Thus, while therapeutic progress is being made in the treatment of AD, breakthroughs in treating the devastating neuroinflammatory component are urgently needed as the number of AD patients rises rapidly worldwide.

Laromestrocel is an allogeneic bone marrow-derived medicinal mesenchymal stem cell (MSC) therapy that has neuromodulatory properties^24–27^. In the Phase 2a CLEAR-MIND study laromestrocel showed clinical efficacy on the Composite Alzheimer’s Disease Score (CADS), Montreal Cognitive Assessment (MoCA) Alzheimer’s Disease Cooperative Scale-Activities of Daily Living (ADCS-ADL) and Mini Mental State Exam (MMSE) and also compelling evidence of reduced brain atrophy in core AD brain regions: hippocampus, temporal lobe, and other brain regions^24^. In that study, there were initial indications of reduced neuroinflammation in the cingulate cortex as assessed through mean diffusivity via diffusion tensor imaging (DTI), and we identified sTIE2 as a potential serum biomarker of AD representing impaired vascular and inflammatory status that was improved with laromestrocel treatment. Here, we investigate brain neuroinflammatory status using free water measurements from the MRI data of participants in the CLEAR-MIND trial. Free water has recently become recognized as a reliable and non-invasive indicator of neuroinflammation in the AD brain, increasing in the hippocampus and other core AD brain regions during the course of the disease^28–31^, and as such holds promise as a technological advancement in AD clinical trial research. Our results indicate widespread abolition of neuroinflammatory drive in the regions of core pathology in the AD brain after treatment with laromestrocel and provide immunomodulatory correlates consistent with peripheral immune system engagement.

## Results

The analysis conducted here was a post hoc assessment of MRI data and plasma biospecimens collected during the 39-week, randomized Phase 2a CLEAR-MIND trial (ClinicalTrials.gov: NCT05233774)^24^. Here we evaluated single-shell free water fraction MRI data from both gray and white matter regions using Freesurfer and Johns Hopkins University (JHU) region maps^32,33^. Multi-modal data analysis using machine learning and deep learning techniques provided new insights on mechanism of action. Treatment groups were: placebo (four monthly doses), 25 million cells (single dose followed by 3 monthly doses of placebo; 25Mx1), 25 million cells (four monthly doses; 25Mx4), and 100 million cells (four monthly doses; 100Mx4)^24^.

### Abrogation of neuroinflammation in core AD brain regions

Analyses of MRI data revealed a progressive increase in neuroinflammation as assessed by free water in the brains of AD patients receiving placebo (N=9), preferentially affecting the hippocampus (p=0.008), temporal cortex (p=0.017) and medulla (p=0.008), over 39 weeks (**Figure 1**). In participants randomly assigned to treatment with laromestrocel, the increases in free water were abrogated at week 39 compared with placebo. In the hippocampus, even low single dose administration of laromestrocel inhibited free water accumulation, whereas in other areas reduced free water was only apparent with repeat dosing: (left hippocampus: 25Mx1: p=0.003; N=10; 25Mx4: p<0.001; N=11; 100Mx4: p<0.001; N=10; left temporal cortex: 25Mx4: p=0.007; N=11; medulla: 25Mx4: p=0.007; N=11; 100Mx4: p=0.007; N=10. **Figure 1**). Within-group comparisons confirmed that free water did not change significantly from baseline to week 39 in any laromestrocel treatment group in left hippocampus (25Mx1: p=0.493, N=10; 25Mx4: p=0.768, N=11; 100Mx4: p=0.974, N=10), left temporal cortex (25Mx1: p=0.291, N=10; 25Mx4: p=0.329, N=11; 100Mx4: p=0.444, N=10), or medulla (25Mx1: p=0.855, N=10; 25Mx4: p=0.672, N=11; 100Mx4: p=0.605, N=10) (**Figure 1**). Crucially, for these regions, the data indicate that inflammation is stabilized with laromestrocel treatment, while progressive inflammation continues in the placebo group. The frontal lobe did not show increased free water in the placebo group, consistent with delayed progression of AD pathology to that area. Like the hippocampus and temporal cortex, the medulla is also an early AD-associated region contributing to autonomic dysfunction characteristics of the disease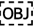^34,35^. The change in free water with respect to placebo is represented on a brain surface map with lateral and coronal views, according to brain region (**Figure 2A, B**).

**Figure 1.**
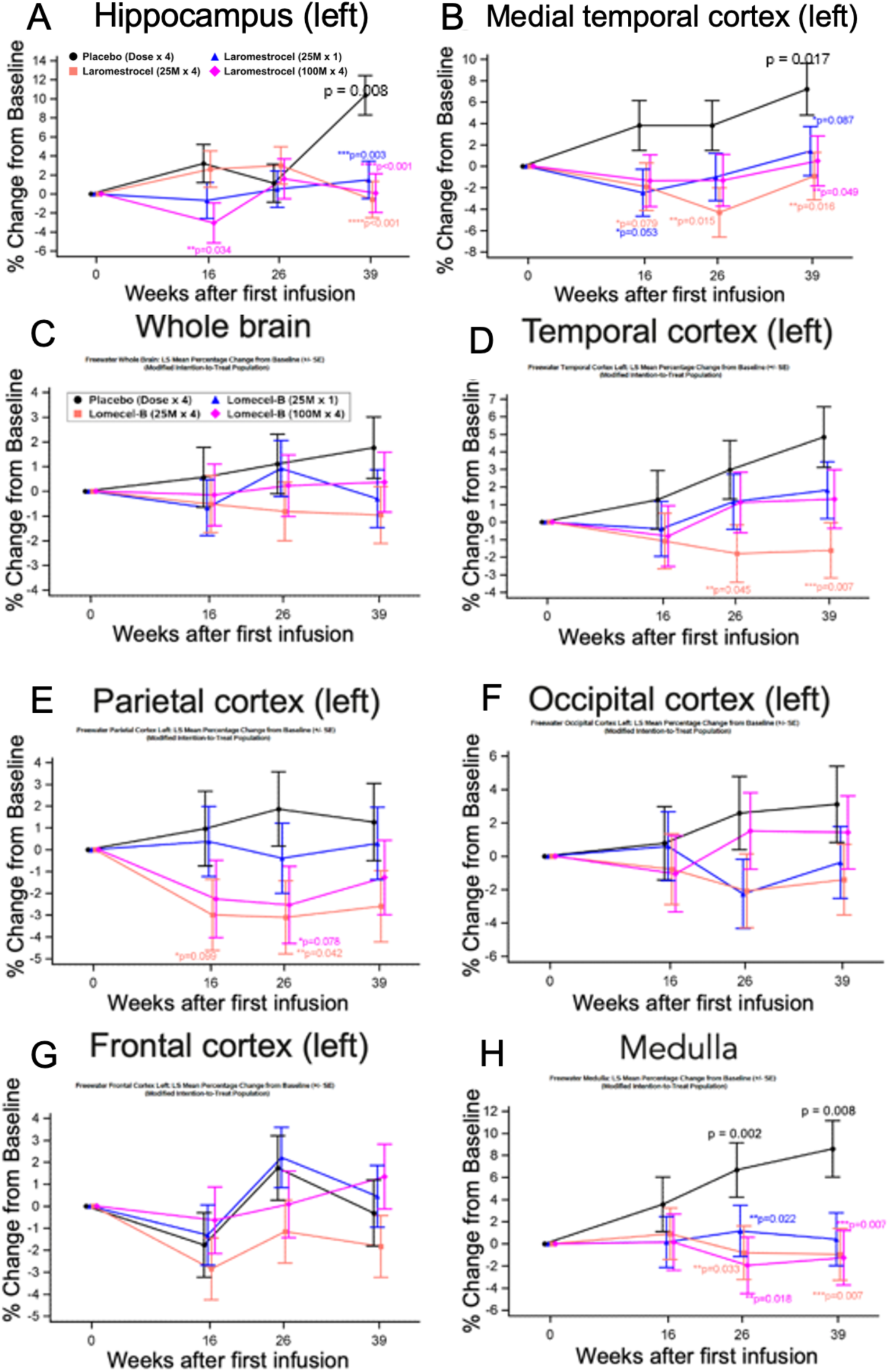
Reduced neuroinflammation in key AD brain regions after laromestrocel treatment. Left hippocampal free water increased by ∼10% in the placebo group (black line) during the 39 week trial period (p=0.008 compared with baseline), while treatment groups showed no increase, reaching statistically significant differences with respect to placebo for all treatment groups (25Mx1: p=0.003, N=10; 25Mx4: p<0.001, N=11; 100Mx4, p<0.001, N=10 (**A**). Similarly, medial temporal cortex free water rose in placebo compared with baseline (p=0.017), while laromestrocel stabilized free water (25Mx4: 0.016, N=11; 100Mx4: p=0.049, N=10) compared with placebo (**B**). Whole brain free water numerically increased by ∼2% in the placebo group (black line) during the 39-week trial period, while treatment groups showed no increase (**C**). Similar findings for temporal, parietal, and occipital lobes showed statistical significance: Temporal lobe, week 39, 25Mx4: p = 0.007, N=11; (**D**); Parietal lobe, week 26, 25Mx4, p=0.042, N=10 (**E**); Medulla, 25Mx4, week 39, p=0.007, N=11; 100Mx4, week 39, p=0.007, N=10 (**H**), while Occipital cortex showed numeric improvement of all treatment groups (**F**). Frontal cortex did not show a treatment effect (**G**). Statistical comparisons utilized an MMRM model. Error bars represent ± SEM.

**Figure 2.**
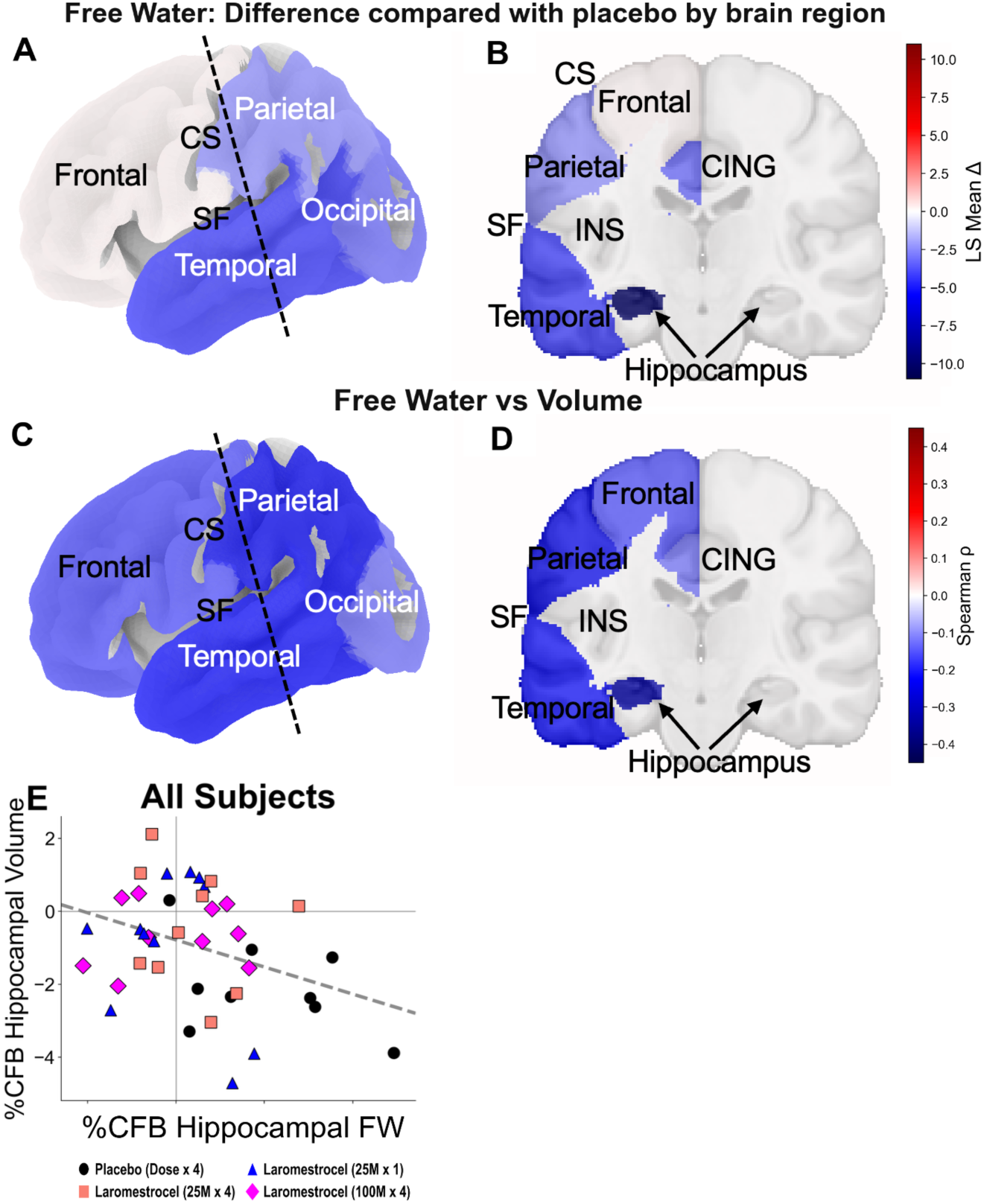
Decreased inflammation and atrophy in core AD brain regions after laromestrocel treatment. Pooled treatment group free water changes (LS mean differences) were mapped onto the brain using the Harvard–Oxford atlas for volumetric regions and the Destrieux atlas for cortical surface parcels, with values displayed using a symmetric seismic (blue, white, and red) colormap. Blue shades represent a negative difference of pooled treatment groups compared with placebo. Lateral (**A**), and coronal section (**B**) views showing the hippocampus (hipp) (**H**). The hippocampus shows the largest anti-inflammatory effect of laromestrocel treatment compared with placebo, followed by the temporal lobe. Spearman correlation analyses at Week 39 across all subjects (N = 40) showed that greater regional free water reduction was associated with greater brain volume preservation, with the strongest inverse correlation in the left hippocampus (ρ = −0.33, p=0.040) and trends in the temporal (ρ = −0.22, p = 0.177) and parietal lobes (ρ = −0.24, p = 0.128), mapped onto brain surfaces (**C**, **D**). Per-subject scatter plot of left hippocampal free water vs hippocampal volume percent change from baseline at Week 39 for all subjects (**E**) and the dashed line indicates the linear regression fit. Dotted lines in A, C indicate the approximate plane of coronal section in B, D. FW, free water; SF, Sylvian Fissure; CING, cingulate cortex; INS, insular cortex; CS, central sulcus.

### Reduced neuroinflammation correlates with increased brain volume preservation

Next, we analyzed whether the lack of free water increase after laromestrocel treatment regionally correlated with reduced brain atrophy. We performed Spearman correlation analyses at 39 weeks for individual brain regions and found an inverse correlation between free water and regional volume, reaching statistical significance for the hippocampus (ρ=-0.326, p=0.040, N=40), but not other areas: temporal (ρ=-0.218, p=0.177, N=40), parietal (ρ=-0.245, p=0.128, N=40), frontal (ρ=-0.155, p =0.338, N=40), and occipital lobes (ρ=-0.126, p=0.439, N=40) (**Figure 2C, D**). The treatment group-wise linear regression plot for the hippocampal free water/volume relationship is shown in **Figure 2E**. These data indicate that reduced neuroinflammation in core AD and other brain regions corresponds with reduced brain atrophy at 39 weeks.

### Reduced neuroinflammation correlates with improved performance on clinical assessments

Our central hypothesis is that laromestrocel-reduced brain inflammation leads to reduced brain tissue destruction and ultimately to improvements in patient cognition, function, and quality of life. In order to test this hypothesis, we correlated free water with key clinical metrics including MoCA, MMSE, ADCS-ADL, where the direction of improvement is a higher score, and the CDR-SB, and ADAS-COG13, where the direction of improvement is a negative score. Spearman correlation analysis at week 39 showed correlations in the direction of improvement in all cases. We overall found *inverse* correlations with the MoCA, MMSE, and ADCS-ADL across brain regions, reaching statistical significance in whole brain for MoCA (ρ= −0.440, p=0.005, N=39), while correlations were overall *positive* across brain regions for the CDR-SB and ADAS-COG13, reaching statistical significance for ADAS-COG13 (ρ= 0.405, p=0.009, N=40) (**Figure 3**). Other comparisons are listed in **Table S1**. Correspondingly, reduced brain atrophy also correlated with improved clinical outcomes, reaching statistical significance for MMSE: ρ=0.411, p=0.007, N=42 and ADCS-ADL: ρ=0.353, p=0.022, N=42, and trending in the direction of improvement for MoCA: ρ=0.280, p=0.076, N=41 (**Figure 4**). Other comparisons are listed in **Table S2**.

**Figure 3.**
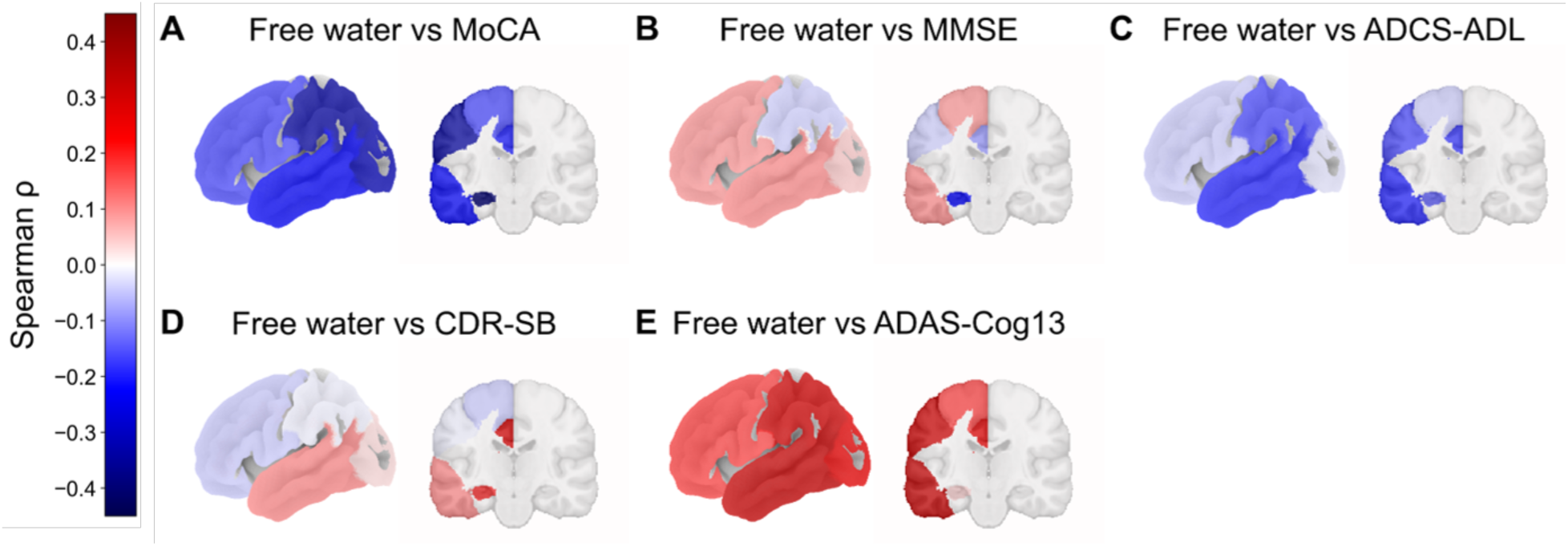
Reduced neuroinflammation correlates with improved clinical outcomes. Left-hemisphere brain surface maps of Spearman correlation coefficients (ρ) between placebo-adjusted percent change from baseline in regional free water and placebo-adjusted percent change from baseline in clinical scores at Week 39: (A) MoCA, (B) MMSE, (C) ADCS-ADL, (D) CDR-SB, and (E) ADAS-Cog13. Cool colors indicate inverse associations; warm colors indicate positive associations (scale shown). For scores where clinical improvement is a positive change (MoCA, MMSE, ADCS-ADL), free water reduction showed mostly inverse correlations. The strongest regional inverse correlations were for free water vs MoCA (left hippocampus: ρ = −0.40, p = 0.011, N = 39), parietal cortex (ρ = −0.35, p = 0.030, N=39) and occipital cortex (ρ = −0.32, p = 0.045, N = 39). For CDR-SB and ADAS-Cog13, where clinical improvement is a negative change, regional correlations were mostly positive, reaching statistical significance for temporal cortex in ADAS-Cog13 (ρ = 0.322, p = 0.043, N = 40). Placebo adjustment reflects change relative to the placebo arm mean prior to correlation analysis.

**Figure 4.**
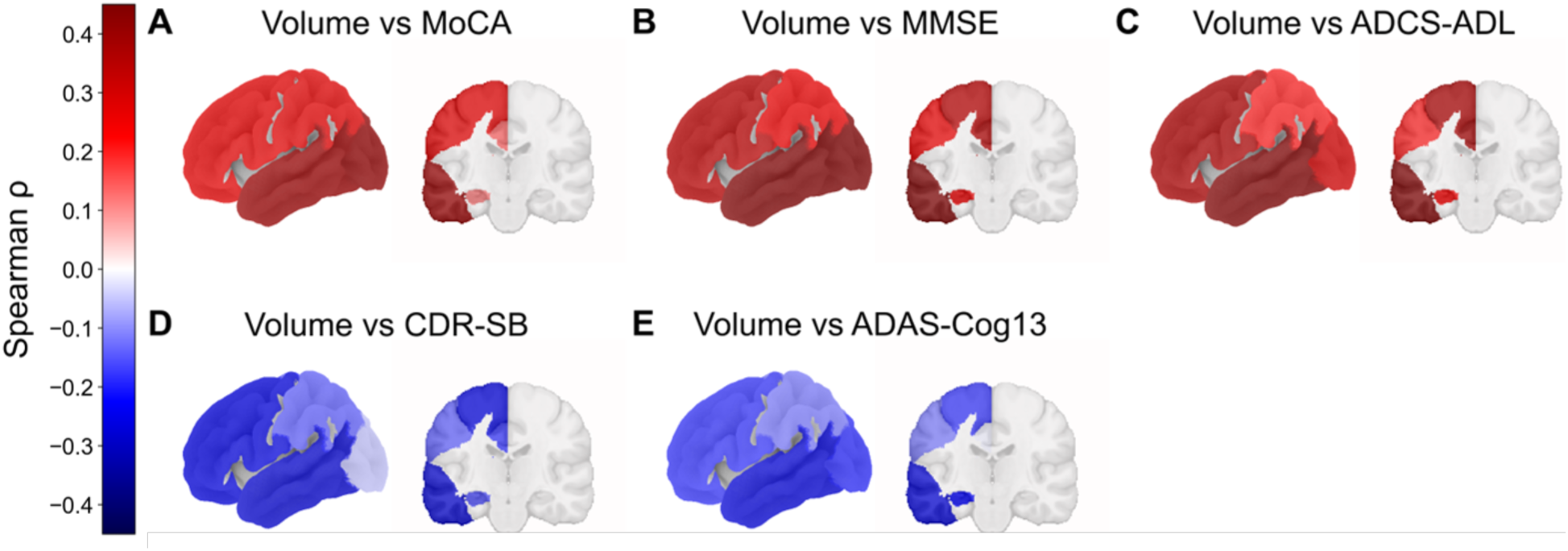
Brain volume preservation correlates with improved clinical outcomes. Left-hemisphere brain surface maps of Spearman correlation coefficients (ρ) between placebo-adjusted percent change from baseline in regional brain volume and placebo-adjusted percent change from baseline in clinical scores at Week 39 (N = 42): (A) MoCA, (B) MMSE, (C) ADCS-ADL, (D) CDR-SB, and (E) ADAS-Cog13. Warm colors indicate that volume preservation correlates with clinical improvement on positive-direction scales; cool colors indicate inverse associations (scale shown). The strongest regional correlations were observed for volume vs MMSE (Frontal Cortex: ρ = 0.35, p = 0.025; Temporal Cortex: ρ = 0.47, p = 0.002; Occipital Cortex: ρ = 0.49, p = 0.001; Cingulate Cortex: ρ = 0.36, p = 0.018, N = 42); MoCA (Temporal Cortex: ρ = 0.40, p = 0.009; Occipital Cortex: ρ = 0.42, p = 0.007, N = 41), and ADCS-ADL (Frontal Cortex: ρ = 0.370, p = 0.016; Temporal Cortex: ρ = 0.46, p = 0.002; Cingulate Cortex: ρ = 0.390, p = 0.011, N= 42). Placebo adjustment reflects change relative to the placebo arm mean prior to correlation analysis.

### Reduced neuroinflammation in key AD white matter tracts

Analysis of free water of brain white matter regions showed similar trends in white matter tracts associated with AD. The fornix, which is a principal white matter tract serving the hippocampus and has been reported to show pathology in AD^29,36^, showed increasing free water over the 39-week trial in the placebo population (**Figure S1a**). All three groups receiving laromestrocel showed numerically reduced free water compared with placebo, although the reduction did not reach statistical significance (25Mx4, week 26: p=0.058, N=10). The sagittal stratum, which is a key crossroads white matter tract mediating communication between the temporal lobe and other lobes and impacted in relation to temporal lobe degeneration in AD^37^, showed a similar trend (100Mx4, week 39: p =0.096, N=10). The uncinate fasciculus, which is another site of early white matter degeneration in AD along with fornix, corpus callosum, and other regions^38–40^, showed a similar trend (100Mx4, week 39: p=0.064, N=10). (**Figure S1**).

### Blood biomarker evidence for reduced neuronal damage and peripheral immune system engagement

Plasma biomarkers for neuronal damage include glial fibrillary acidic protein (GFAP), neurofilament light (NfL), a neuron-specific cytoskeletal protein enriched in axons^41^, and neurogranin, a neuron-specific calmodulin-binding protein enriched in synapses, both of which are reportedly elevated in cerebrospinal fluid (CSF) of AD patients^42^. Rising neurogranin levels in CSF are indicative of synaptic damage and have been correlated with brain atrophy in AD^43–46^, and emerging evidence has indicated a correlation between plasma neurogranin and neurodegeneration (cortical thinning) in AD^47^. We examined GFAP, NfL and neurogranin using the ARGO NuLISA CNS120 assay^48^ and found that neurogranin progressively rises in the placebo group, while all groups receiving laromestrocel showed reduced plasma levels of neurogranin at week 39, reaching statistical significance for 25Mx4 (p=0.023, N=11) (**Figure S2A**), but detected no significant differences compared with placebo for NfL or GFAP. No significant differences in Amyloid β 42/40 ratio via the Lumipulse Fujirebio assay or pTau217 via NuLISA were observed at 39 weeks post treatment versus placebo (**Figure S2b; Figure S2c**).

The lack of change in canonical Amyloid and Tau biomarkers may indicate that laromestrocel does not act primarily through these pathways. To identify candidate mediators in an unbiased manner, we performed an ordinary least squares (OLS) regression screen across 117 proteins on the NuLISA CNS120 panel, comparing pooled active treatment groups to placebo at Week 39 (**Figure 5a)**^49^. Two proteins reached nominal significance, both elevated in the pooled treatment arm: Interleukin-13 (β = +7.52%, p = 0.003) and Interleukin-9 (β = +8.75%, p = 0.025). Increased IL-13 in plasma has been found to engage peripheral immune components that have been linked to the modulation of brain microglia and inflammatory activity^50–53^. Both identified biomarkers are canonical Th2 cytokines associated with anti-inflammatory and regulatory immune programs, providing a first directional clue regarding an immune signature of laromestrocel treatment.

**Figure 5.**
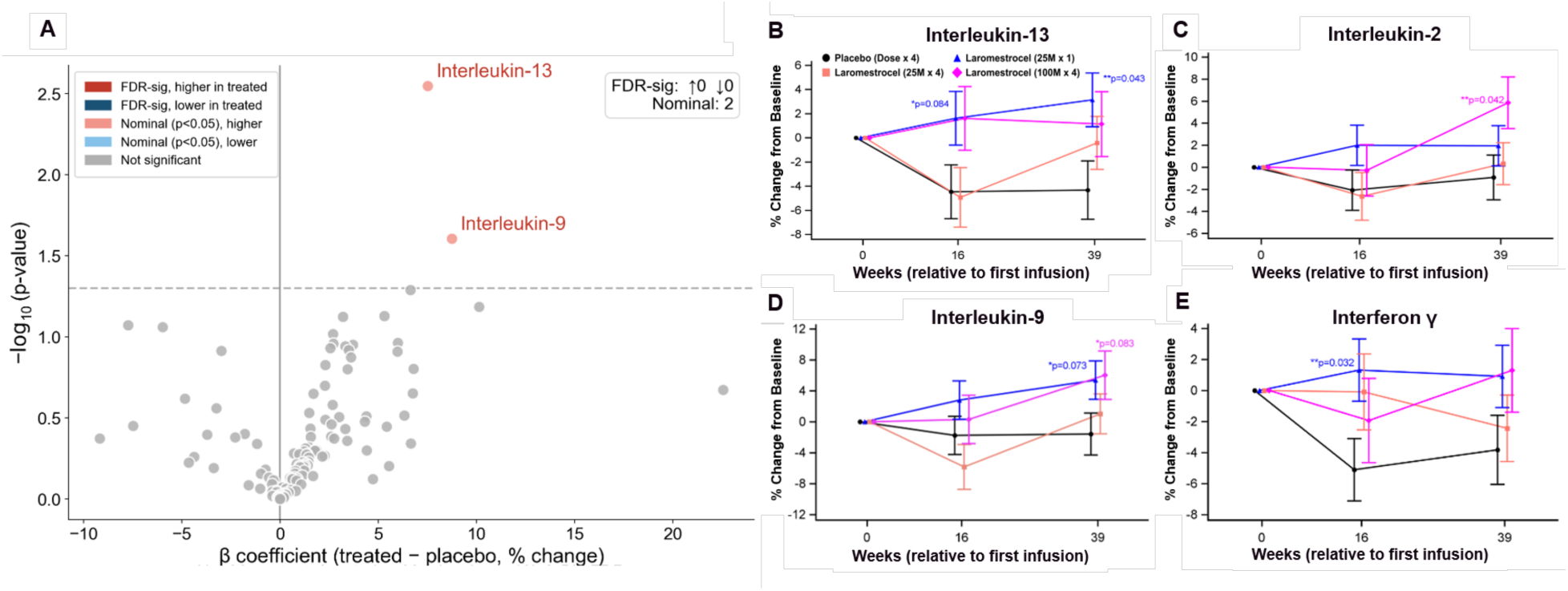
An unbiased plasma protein screen identifies Th2 cytokines as top treatment-responsive signals, with converging peripheral immune activation. (A) Volcano plot of OLS regression β coefficients (treatment minus placebo, % change from baseline at Week 39) versus −log₁₀ nominal p-value across 117 NuLISA CNS120 proteins, adjusted for sex and APOE-ε4 status (N = 36). Two proteins reached nominal significance (dashed line, p = 0.05), both elevated in the treatment arm: Interleukin-13 (β = +7.52%, p = 0.003) and Interleukin-9 (β = + 8.75%, p = 0.025); zero proteins reached FDR significance. (B) IL-13 increased in all treatment groups at Week 39, reaching statistical significance for 25Mx1 (p = 0.043, N = 10). (C) IL-2, a regulator of T-regulatory cell expansion, showed statistically significant elevation at Week 39 (100Mx4: p = 0.042; N=7). (D) IL-9 showed trend-level elevations at Week 39 (25Mx1: p = 0.073, N=10; 100Mx4: p = 0.083; N=7). (E) Interferon-γ was elevated compared with placebo at Week 16, consistent with peripheral immune activation (25Mx1: p = 0.032, N = 10). Statistical comparisons in B–E utilized an MMRM model. Error bars represent ± SEM.

Examination of the broader peripheral immune cluster using MMRM revealed elevations in cytokines associated with Th2 and regulatory immune responses. Analysis of plasma interleukins via NuLISA showed an increase in IL-13 in all treatment groups at week 39, reaching statistical significance for 25Mx1 (p = 0.043, N=10; **Figure 5b**). IL-9, another Th2-associated cytokine, was similarly elevated in treatment groups compared with placebo at week 39, with trend-level increases for 25Mx1: (p=0.073, N=10) and 100Mx4 (p=0.083, N=7; **Figure 5d**). A key regulator of T-regulatory cell expansion, IL-2^54^, was also increased compared with placebo by week 39 (100Mx4: p=0.042, N=7; **Figure 5c**). Increased Interferon-γ, an indicator of peripheral immune activation, was also observed at weeks 16 and 39, reaching statistical significance for 25Mx1 at week 16 (p=0.032; N=10) (**Figure 5e**).

### Hypothesis-free classification confirms coordinated multimodal treatment signature

Finally, to determine if the treatment signal was powerful enough to be detected at the individual patient level, a supervised machine learning analysis was performed on the combined feature set of MRI imaging data (free water) and plasma biomarkers. This test is stricter than group-mean comparisons alone because it requires the treatment signal to be consistent across individual subjects. Random Forest Classifier (**Figure 6**) and Extreme Gradient Boosting Classifier (**Figure S3**) were trained to distinguish treated patients from placebo patients using Week 39 percent-change values of freewater in Freesurfer-map grey matter regions and in JHU-map white matter tracts along with NuLISA plasma proteins. Both models classified patients above chance, with balanced accuracy of 0.683 (95% BCa CI: 0.517–0.867) and AUC of 0.750 (0.550–0.950) for Random Forest, and balanced accuracy of 0.633 (0.345–0.750) and AUC of 0.750 (0.367–0.900) for XGBoost. Performance of this magnitude in a 43-patient dataset, where classifiers are prone to noise and overfitting, indicates that the multimodal treatment signature is detectable across individuals without prior specification of a predictive feature set. As a negative control, identical pipelines applied to Week 0 baseline values yielded near-chance performance (**Figure S4**), confirming that classifier accuracy reflects post-treatment biology and is unlikely to be driven by pre-existing group differences.

**Figure 6.**
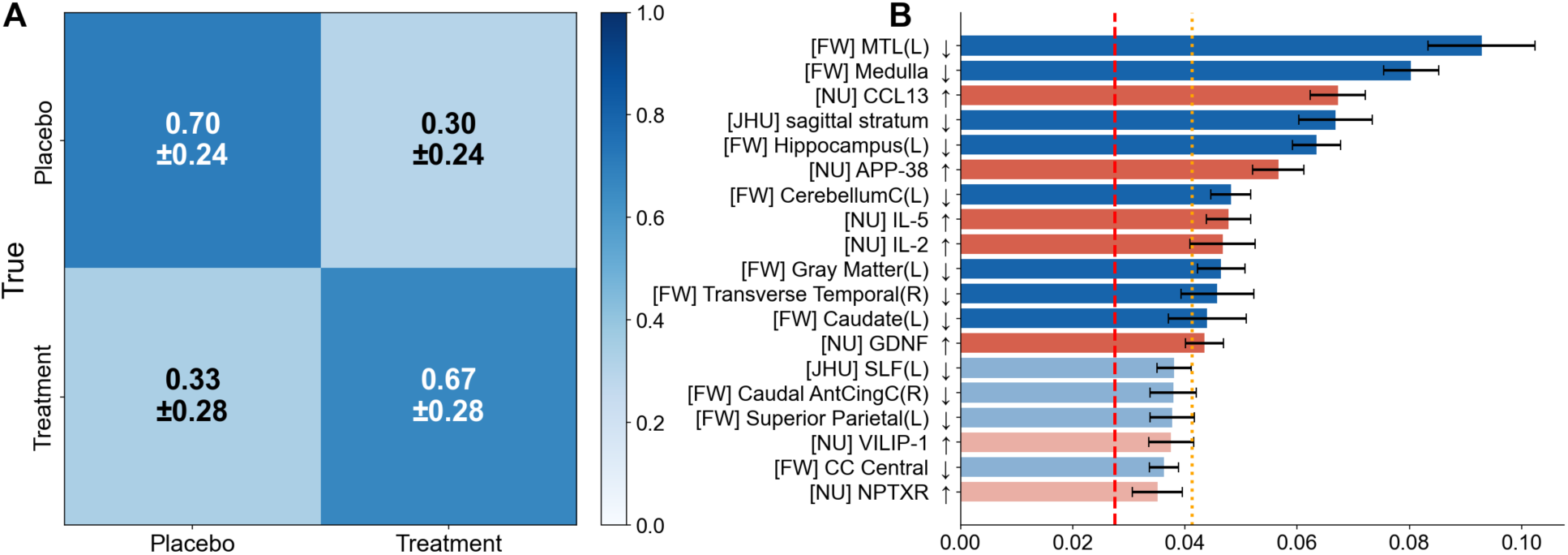
Multimodal machine learning analysis identifies free water in core AD brain regions and immunomodulatory biomarkers as top predictors of treatment. Classification of treatment vs. placebo at Week 39 used free-water diffusion MRI (FW), JHU white matter tract metrics, and NuLISA plasma protein biomarkers (N=43). Random Forest (RF) classifier was trained on a stabilized feature set of 19 features selected by majority vote across screening runs from 289 total features. **(A)** RF confusion matrix: 70% of placebo and 67% of treatment subjects were correctly classified (balanced accuracy [BACC] = 0.683 [95% BCa CI: 0.517– 0.867]; AUC = 0.750 [0.550–0.950]). **(B)** The 19 features ranked by mean RF importance across folds. Bar color encodes the univariate direction of each feature’s Week 39 percent-change contrast (treatment minus placebo): red bars indicate features that increased in the treatment group relative to placebo (e.g., plasma cytokines), while blue bars indicate features that decreased (e.g., regional free-water, reflecting reduced neuroinflammation); color intensity distinguishes features exceeding 1.5× the shadow threshold (fully saturated) from those exceeding the shadow threshold but not the 1.5× margin (half-saturated). Direction is a descriptive marginal summary and does not isolate individual feature contributions within the multivariate classifier. Gray bars indicate features at or below the shadow noise floor. Red dashed lines indicate mean shadow-feature importance; orange dotted lines mark the 1.5× shadow retention threshold. Error bars denote SEM across folds. MTL: medial temporal lobe, CC, corpus callosum.

Top predictive features included free-water in canonical AD-affected regions such as medial temporal lobe, hippocampus, medulla, temporal cortex, caudate, and certain white matter tracts including the sagittal stratum and superior longitudinal fasciculus, and plasma markers including CCL13, IL-2, IL-5, and GDNF. While the classifiers themselves do not assign directionality, cross-referencing the feature list with the univariate analyses revealed a consistent pattern: in the treatment arm, all predictive free water features decreased while all predictive plasma markers increased. The co-movement of the reduction in regional neuroinflammation with the increase in top plasma biomarkers enriched in anti-inflammatory immune engagement suggests a coordinated multimodal response. The hypothesis-free recovery of both the core AD-associated brain regions and the peripheral immune axis identified by directed analyses supports the interpretation that laromestrocel produces a biologically coherent and detectable treatment effect.

## Discussion

Using free water MRI analysis of patients with mild AD^24^, here we show that laromestrocel mitigates the progressive brain neuroinflammation evident in AD. Systemic infusions of laromestrocel, an MSC with medicinal paracrine effects, produce a potent blockade of neuroinflammatory progression in the brain, in a manner associated with preservation of brain volume and improvements in cognitive, functional, and quality of life measures. Collectively, these results support an anti-inflammatory mechanism of action of laromestrocel that directly preserves and, in some cases, reverses brain atrophy associated with AD. Further, these phenotypic observations are directly associated with clinical benefits and support ongoing clinical development.

These results support a central hypothesis in AD pathophysiology: that mitigating neuroinflammatory drive in the AD brain reduces neuronal damage and death, leading to less brain tissue destruction and improved cognitive and functional outcomes. The apparent ability of laromestrocel to durably interrupt this disease mechanism indicates that laromestrocel is potentially a highly significant contribution to the armamentarium of therapies that can be leveraged in AD treatment. Laromestrocel did not appear to acutely influence Amyloid or Tau levels in plasma, although we cannot rule out a benefit to the blood brain barrier and amyloid export, which is known to require a longer time scale to show changes in plasma^55^. This will be tested in a future, longer trial.

### Limitations of the study

This was an exploratory, post hoc analysis of a trial of relatively short duration and small sample size (n=49), which was not powered on the basis of free water or blood biomarker endpoints. Given the small sample size, it is not surprising that a consistent dose response was not observed, as indicated in the parent report of the CLEAR MIND trial^24^, but nonetheless the principal effect observed in the hippocampus was more pronounced with repeat dosing. Results are therefore hypothesis generating as they are without correction for multiple comparisons. Additionally, free water is not a pure metric of neuroinflammation, as it can also reflect changes in blood flow or blood brain barrier (BBB) dynamics that are not deciphered here. The machine-learning classifiers were trained and evaluated on the same small, high-dimensional dataset used for univariate analyses, precluding fully independent validation and increasing susceptibility to overfitting. Although there was evidence of reduced plasma neurogranin, the unchanged NfL and GFAP in blood plasma was unexpected given the preservation of brain tissue volume in subjects receiving laromestrocel; future trials will assess NfL and GFAP in CSF. Additionally, we emphasize that the molecular immunomodulation sequence posited here is hypothesis-generating: we did not directly measure Treg frequency, IDO activity, or MSC licensing, which will be directly tested in a future trial with greater statistical power.

### How might laromestrocel reduce brain neuroinflammation?

Firstly, although other mesenchymal stem cell therapies have been shown not to cross the healthy BBB in large numbers^56^, being sequestered in the lung and other organs^57^, it is unknown whether laromestrocel may cross into the brain of AD patients. Studies have demonstrated increased macrophage infiltration of the AD brain due to a compromised BBB^58,59^, and we speculate laromestrocel could, too, facilitating paracrine signaling mechanisms in the brain parenchyma. This will be examined in a future study. Alternatively, laromestrocel may engage the peripheral immune system and indirectly influence brain microglial activity and inflammatory drive. Here we present limited support for this hypothesis: increased IL-13, reported to reduce microglial hyper-inflammatory activity^50–53^, and general peripheral immune engagement through increased Interferon-γ. IFN-γ is a recognized licensing cue for MSC immunomodulation, and IFN-γ-primed MSCs have been shown to promote regulatory T-cell (Treg) induction and immunosuppressive macrophage polarization^60^. MSC efferocytosis by peripheral macrophages could also feature in this process^61^, or as reported previously, through engagement of Treg cells that feed back to regulate brain microglial activity^62^. Our observation of increased IL-2, a key driver of Treg expansion^54^, in plasma is consistent with treatment-emergent T-regulatory cell activation and with an immunomodulatory mechanism of action of MSCs. This key observation supports a peripheral-to-central mechanism of action reportedly able to reduce microglial pro-inflammatory properties.

A third, complementary mechanism may involve preservation of vascular and blood– brain barrier (BBB) integrity through the angiopoietin–TIE2 axis, whose disruption sensitizes the vascular endothelium to TNF-α and promotes inflammation^63^. Soluble TIE2 (sTIE2), shed from the endothelial receptor, may reflect this disruption. In CLEAR-MIND, sTIE2 rose progressively in the placebo group (p = 0.047 at week 26) but was significantly reduced by laromestrocel^24^, and a dose-dependent reduction was independently replicated in our Phase 2b frailty trial^27^.

Because BBB leakage could contribute both to neuroinflammation and to the extracellular free water signal, restored TIE2 signaling offers a mechanistically convergent explanation, although sTIE2 is a peripheral measure that does not by itself establish a CNS mechanism.

Such cellular and molecular mechanisms should continue to be explored to determine how laromestrocel may exert a sustained anti-inflammatory effect. An exciting possibility is that laromestrocel may reprogram immune regulatory systems. Such durable and transformative effects are not often seen with small molecule therapies that acutely target single enzymes of pathways. The pleiotropic nature of stem cell therapies offers new mechanisms of action and potentially more comprehensive treatment of complex disease states that warrants further investigation. We suggest that laromestrocel, as a long-lasting anti-inflammatory therapy, could be paired with anti-Amyloid immunotherapies for a syngergistic therapeutic benefit in AD treatment, potentially limiting the risk of ARIA observed with the anti-amyloid therapies while directly addressing both AD inflammation and Amyloid accumulation.

## Materials and Methods

The CLEAR-MIND study (ClinicalTrials.gov: NCT05233774) was a 49-patient, randomized, placebo controlled, proof-of-concept study that tested 3 different dosing regimens of intravenous (IV) delivery of laromestrocel vs placebo in patients with mild AD dementia^24^. The design explicitly tested whether repeated dosing of laromestrocel provided added clinical benefits compared with single dosing. The treatment groups were: four monthly intravenous (IV) infusions of Placebo (*N*=12), 25 million (25M) cells once followed by 3 monthly infusions of placebo (25Mx1; *N*=13); 25 million (25M) cells monthly for 4 months (25Mx4; *N*=13), or 100 million (100M) cells monthly for 4 months (100Mx4; *N*=11)^24^. Patients were longitudinally assessed for cognitive, functional, quality of life, and volumetric MRI and DTI imagery until the end of the trial at week 39 as described previously^24^.

### Free Water

We extracted free water fraction data from the CLEAR-MIND MRI DTI data from nearly 150 brain regions. Longitudinal data were collected using 3T MRI imagery at baseline and weeks 16, 26, and 39^24^. Diffusion MRI images were modeled using a single shell bitensor model that separates isotropic (free water) and anisotropic (tissue) diffusion components^64^. Brain sub-regions were grouped into broad categories representing the major brain lobes and circuit systems, e.g. temporal, parietal, occipital, frontal, limbic, hippocampal etc. We focused on the core AD-associated brain regions for pathogenesis, particularly the hippocampal and temporal regions, which include the entorhinal cortex, and which are among the first brain regions to show AD pathology and atrophy. Raw free water values represented the voxel-wise fraction of free water. The master dataset was quality-controlled independently by TruMinds and certified.

### Plasma Biomarkers

Plasma samples were collected in a longitudinal series of patient visits and sent to Longeveron’s laboratory in Miami, FL where they were kept frozen at −80°C until analysis. Samples were sent to Banner Health (Sun City, AZ) for analysis using the Lumipulse Fijirebio assay for Amyloid beta 42/40 and pTau217, as well as a targeted proteomic analysis: the Alamar ARGO NUcleic acid-Linked Immuno-Sandwich Assay (NuLISA) CNS120 panel^48^. The same plasma sample was assessed at the same time for both Lumipulse and NuLISA assays.

### Statistical processing

All statistical analyses utilized the CLEAR-MIND modified intention-to-treat population. Brain volume, free water, and plasma biomarker endpoints were evaluated group-wise using a mixed model for repeated measures (MMRM) using SAS version 9.4. Least squares mean differences between each active treatment group and placebo were estimated and formally tested against placebo or baseline using a two-sided Wald test. The p-values shown in the figures correspond to these tests. No adjustments for multiple comparisons were applied. Statistical significance was considered met if p < 0.05. Error bars represent SEM.

### Atlas-Based Brain Region Visualization

Three-dimensional brain visualizations for all figures were generated using the nilearn Python package^65^ through a shared pipeline, with figures differing only in the per-region values projected onto the anatomical framework. Cortical surface renderings were produced on the fsaverage left hemisphere using the Destrieux parcellation^66^, while volumetric coronal slices (y = −20 mm) were overlaid on the MNI152 template^67^ and clipped to the MNI152 brain mask to prevent signal bleed beyond the cortical boundary. Cortical lobes were mapped using the Harvard–Oxford cortical atlas and subcortical structures (e.g., hippocampus) using the Harvard–Oxford subcortical atlas (both maxprob, thr0, 1 mm resolution) ^68–72^. Study ROIs were linked to atlas parcels through anatomical keyword matching, and all maps used a symmetric seismic colormap with a fixed scale to allow direct visual comparison across figures.

### Hippocampal Distribution Methods

To visualize the subject-level relationship between hippocampal neuroinflammation and hippocampal atrophy at Week 39, per-subject scatter plots were generated using Week 39 percent change from baseline values for left hippocampal free-water fraction and left hippocampal volume. Three panels were produced: all subjects colored by treatment group, placebo subjects only, and pooled active-treatment subjects only. A linear regression line fitted to each panel’s data is shown alongside reference lines at zero percent change on both axes. Spearman rank correlation coefficients, nominal p-values, and sample sizes are annotated on each panel. All panels share identical axis limits to allow direct visual comparison.

### Volcano Plot Methods

To identify NuLISA proteins whose Week 39 percent change from baseline differed between active-treatment and placebo arms in an unbiased manner, an OLS regression screen was performed across all 117 proteins on the CNS120 panel. Analyses were restricted to participants with paired Week 0 and Week 39 measurements (n = 36; 28 active-treatment subjects across three pooled dose arms, with 8 receiving placebo). For each protein, the model

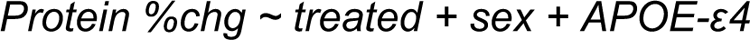

was fitted via OLS, with the binary treatment indicator as the focal predictor and sex and APOE-ε4 carrier status as covariates. Regression coefficients, 95% confidence intervals, nominal p-values, and Benjamini–Hochberg false discovery rate (BH-FDR)-adjusted q-values were extracted for each protein and visualized in volcano-plot form (β coefficient on the x-axis, −log₁₀ nominal p-value on the y-axis) to identify proteins elevated or reduced in the treatment arm relative to placebo.

### Machine Learning Classification of Treatment Response

To evaluate whether post-treatment changes in imaging and plasma biomarkers could distinguish laromestrocel-treated from placebo-treated patients, supervised classifiers were trained to separate active-treatment subjects (pooled; n=33) from placebo subjects (n=10) using Week 39 percent change from baseline values as input features. Classification was performed on a combined dataset of free-water fraction across brain Freesurfer gray matter regions and JHU white matter tract measures, and NuLISA plasma proteins. Features representing bilateral measurements and features with over 40% missingness were excluded, yielding 281 candidate features for the multimodal analysis. Two complementary classifiers were trained. First, a Random Forest (Breiman, 2001) was implemented in scikit-learn^65^ using 300 trees, the square root of the total feature count as the maximum number of features considered at each split, and class-balanced sample weighting. Second, XGBoost (Chen and Guestrin, 2016) was implemented in the XGBoost Python package using 600 boosting rounds, a learning rate of 0.03, a maximum tree depth of 4, and subsample and column-subsample ratios of 0.8 to reduce overfitting.

All reported metrics were derived from a 5-fold cross-validation scheme constructed around the smaller placebo arm, in which placebo subjects were partitioned into five test folds and treatment subjects were randomly sampled within each arm to match placebo counts per fold. Within each fold, feature values were winsorized using six times the median absolute deviation of the training partition to limit outlier influence, and remaining missing values were median imputed from the training partition. Predicted probabilities were averaged across ten random-seed refits per fold before class assignment. To stabilize feature selection, we employed a shadow-feature null reference (Kursa and Rudnicki, 2010), a randomly generated Gaussian noise feature added alongside real features, and retained only features whose importance exceeded 1.5 times the shadow mean in at least 9 of 10 independent screening runs in both classifiers. Model performance was quantified using balanced accuracy and area under the ROC curve (AUC), with 95% bias-corrected accelerated (BCa) bootstrap confidence intervals computed across fold-level estimates (2,000 resamples) (Efron, 1987). As a negative control, identical pipelines were run on Week 0 baseline values, where no treatment effect should exist; this ensured that reported Week 39 performance reflected genuine post-treatment separation rather than random or demographic artifacts. Feature importances were extracted as mean decrease in impurity for Random Forest and gain-based split importance for XGBoost, and averaged across all cross-validation folds and refits to produce stable per-feature rankings; standard errors of the mean were computed across fold-level estimates to quantify uncertainty in each feature’s contribution.

## Data Availability

Please send all requests for data access to the corresponding authors, BGR or JMH. The minimum dataset, without individual patient data, used for the primary, secondary, and exploratory conclusions, may be shared under a data use agreement for IRB approved research. Requests will be considered and responded to within 1 month of receipt. The trial protocol and statistical analysis plan under US FDA IND # 16524 can be shared upon academic or research request.

## Acknowledgements

We thank the Alzheimer’s Association for funding the phase 1 clinical trial and the phase 2a CLEAR-MIND trial^24^, with Part the Cloud initiative grants (PTC C-16-422443; PTC CS-19-623225), Clario for performing the free water MRI data extraction, and Banner Health for performing NuLISA and Lumipulse Fujirebio sample analysis, and Meghan Travis for plasma sample blinding and shipping. We thank the members of the Longeveron team who contributed to the conduct of the original Phase 2a CLEAR-MIND trial^24^.

## Author Contributions

BGR, JMH, and SL wrote the paper and analyzed data. BGR, JB, and SL created figures. JB performed MMRM statistical modeling of free water, Lumipulse, and NuLISA data. SL performed brain map visualization coding, correlation analyses, machine learning analyses, and MMRM analysis. CC, and MI performed free water extraction from MRI datasets. BGR and JMH supervised the project.

---

**Figure S1.**
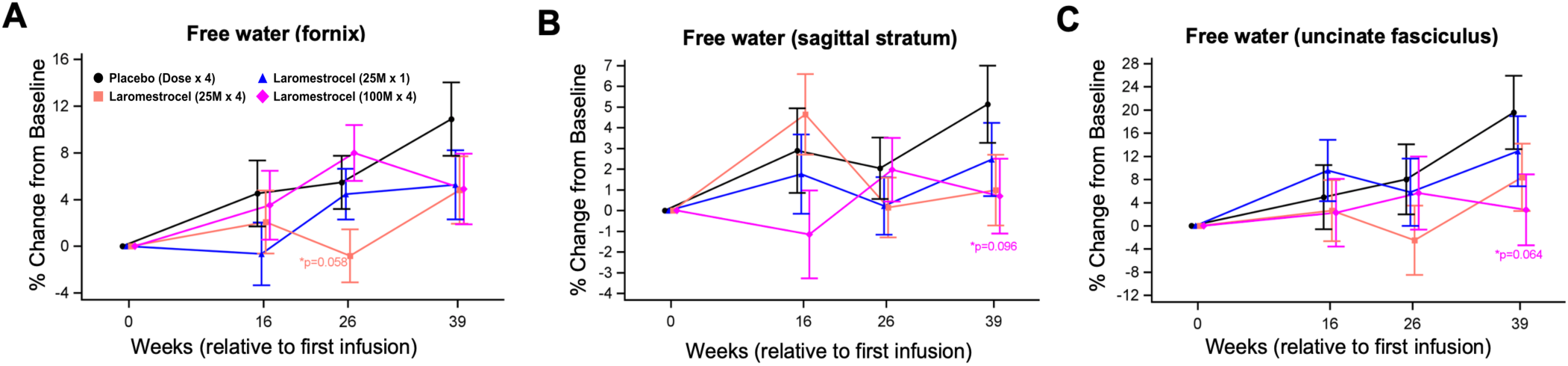
Free water in key AD white matter tracts at week 39. Free water in the fornix steadily increased by more than 10% in the placebo group (black line), while treatment groups showed consistent trends for less increase at week 39 (**A**). Similar trends were found for sagittal stratum (**B**) and uncinate fasciculus (**C**); Statistical comparisons utilized an MMRM model. Error bars represent ± SEM.

**Figure S2.**
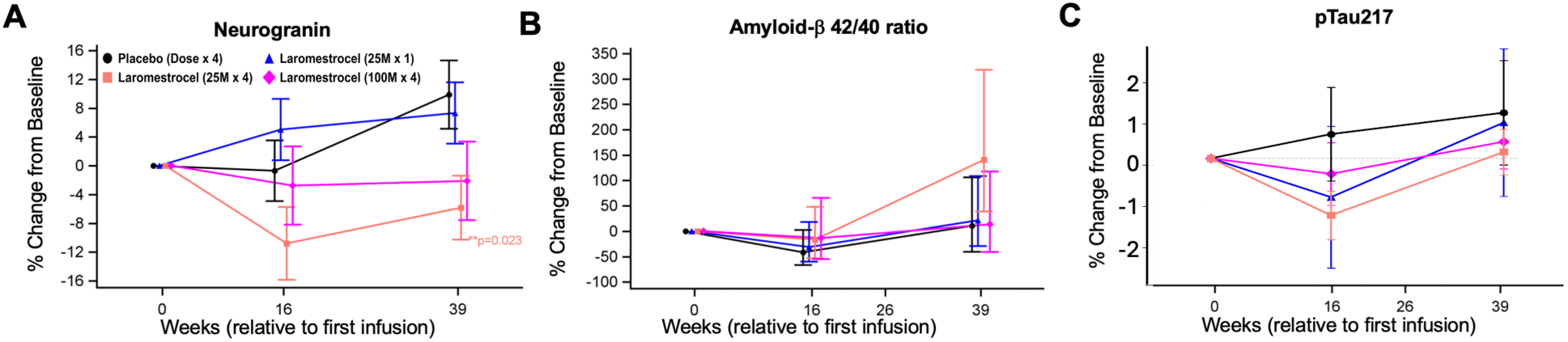
Laromestrocel decreases plasma neurogranin without affecting Amyloid or Tau in plasma. (**A**) Neurogranin, assessed using NuLISA (Banner Health), was reduced compared with placebo, reaching statistical significance for repeat dosing 25Mx4 at week 39 (p=0.023, N=11). (**B**) Plasma Amyloid-b 42/40 ratio and (**C**) pTau217 assessed using the Lumipulse Fujirebio assay and NuLISA, respectively, (Banner Health) showed no significant trends differing from placebo over the course of the 39-week trial. Statistical comparisons utilized an MMRM model. Error bars represent ± SEM.

**Figure S3.**
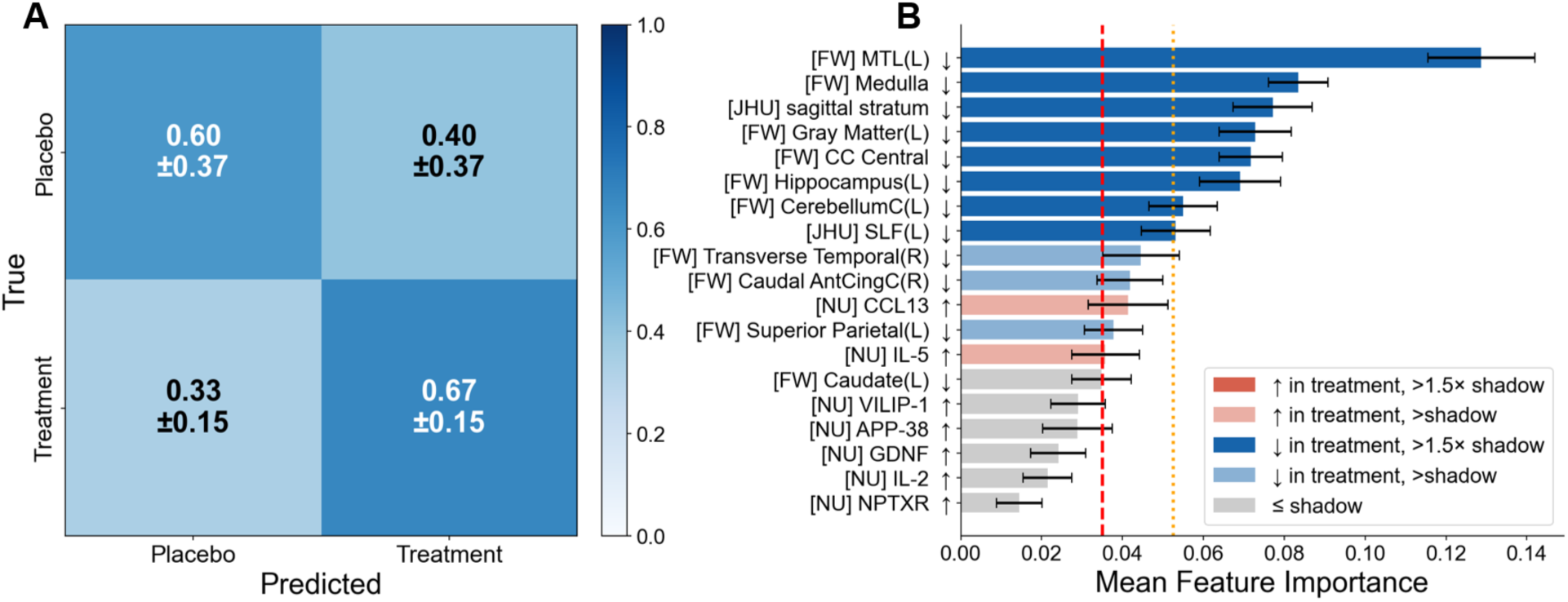
Additional machine learning analysis using a second model confirmed free water in core AD brain regions and immunomodulatory biomarkers as top predictors of treatment. Extreme Gradient Boosting (XGB) classifiers were trained on a stabilized feature set of 19 features selected by majority vote across screening runs from 289 total features (N=43). **(A)** XGB confusion matrix: 60% of placebo and 67% of treatment subjects were correctly classified (BACC = 0.633 [0.345–0.750]; AUC = 0.750 [0.367–0.900]). **(B)** The 19 features ranked by mean XGB importance across folds. Bar color encodes the univariate direction of each feature’s Week 39 percent-change contrast (treatment minus placebo): red bars indicate features that increased in the treatment group relative to placebo (e.g., plasma cytokines), while blue bars indicate features that decreased (e.g., regional free-water, reflecting reduced neuroinflammation); color intensity distinguishes features exceeding 1.5× the shadow threshold (fully saturated) from those exceeding the shadow threshold but not the 1.5× margin (half-saturated). Direction is a descriptive marginal summary and does not isolate individual feature contributions within the multivariate classifier. Gray bars indicate features at or below the shadow noise floor. Red dashed lines indicate mean shadow-feature importance; orange dotted lines mark the 1.5× shadow retention threshold. Error bars denote SEM across folds.

**Figure S4.**
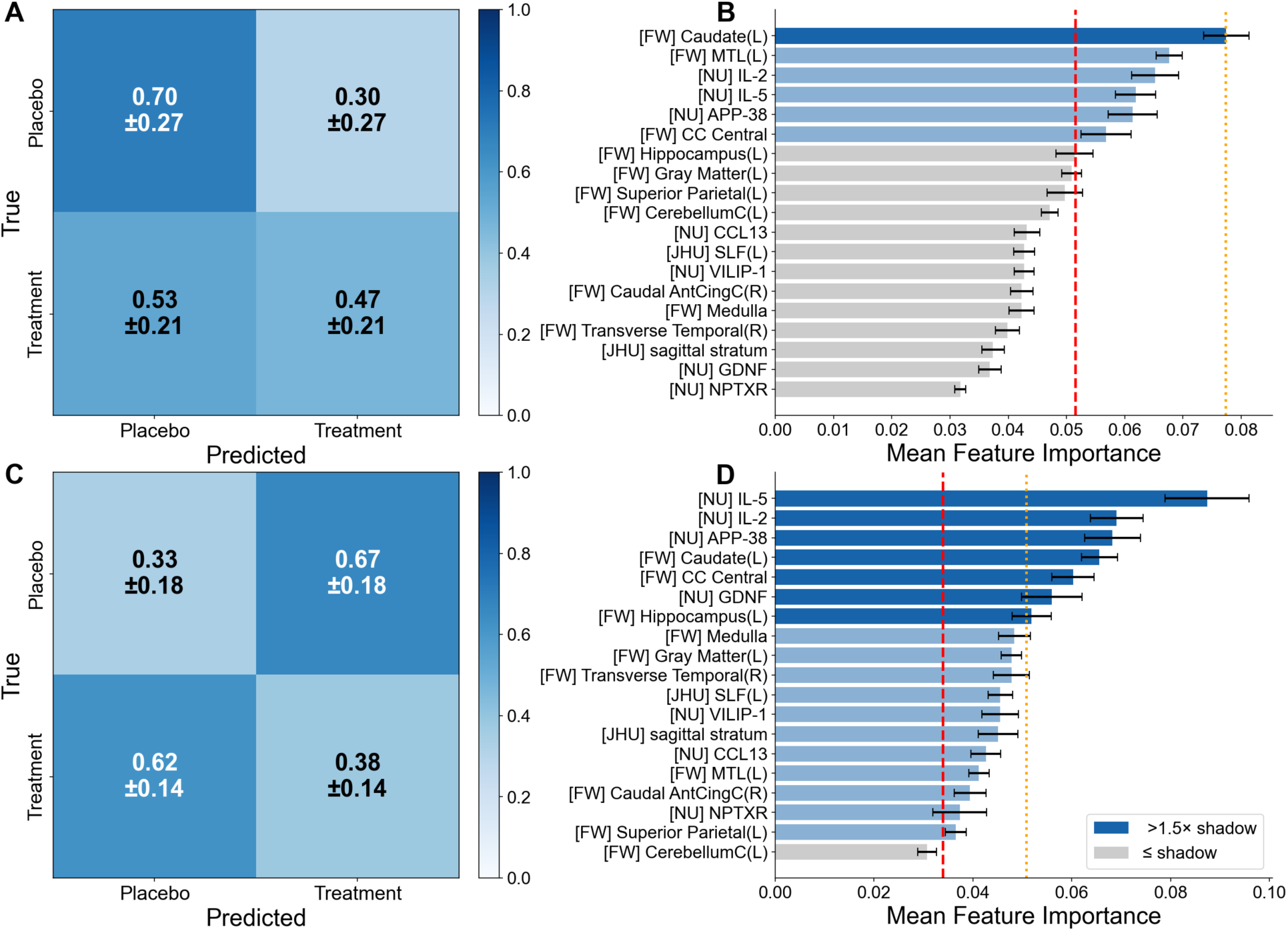
Classifier performance at Week 0 baseline confirms that Week 39 treatment signal reflects post-treatment biology. RF and XGB classifiers with identical pipelines to Figure 7 were applied to Week 0 baseline values across the multimodal feature set, where no treatment effect should exist. (A) RF Week 0 confusion matrix: BACC = 0.583 [0.422–0.717]; AUC = 0.628[0.513–0.731]. (B) Top 20 features by mean RF importance at Week 0. (C) XGB Week 0 confusion matrix: BACC = 0.356 [0.222–0.422]; AUC = 0.454 [0.289–0.593]. (D) Top 20 features by mean XGB importance at Week 0. Performance at Week 0 is substantially attenuated compared with Week 39 (Figure 7), and Week 0 feature importance profiles lack the directional structure seen post-treatment (no separation of decreasing imaging features from increasing plasma markers), consistent with Week 39 classifier performance reflecting a treatment signal rather than pre-existing group differences. Error bars denote 95% BCa bootstrap confidence intervals across cross-validation folds.

**Table S1.** Spearman correlation coefficients and p-values between placebo-adjusted free water percent change and placebo-adjusted clinical score percent change at Week 39, reported for seven brain regions of interest and their cross-region mean (N = 42, except MoCA: N =41).

| Table S1 |  |  |  |  |  |
| --- | --- | --- | --- | --- | --- |
|  | **ADAS-Cog13**<br>(Rho, p) | **ADCS-ADL**<br>(Rho, p) | **CDR-SB**<br>(Rho, p) | **MMSE**<br>(Rho, p) | **MoCA**<br>(Rho, p) |
| **Frontal Cortex (Left)** | 0.165,<br>0.308 | -0.042, 0.798 | -0.043, 0.794 | 0.088, 0.587 | -0.158, 0.335 |
| **Hippocampus (Left)** | 0.039,<br>0.813 | -0.140, 0.389 | 0.181, 0.263 | -0.220, 0.172 | -0.401, 0.011 |
| **Temporal Cortex (Left)** | 0.322,<br>0.043 | -0.194, 0.229 | 0.103, 0.520 | 0.099, 0.545 | -0.229, 0.160 |
| **Parietal Cortex (Left)** | 0.306,<br>0.055 | -0.161, 0.320 | -0.013, 0.934 | -0.042, 0.797 | -0.348, 0.030 |
| **Occipital Cortex (Left)** | 0.259,<br>0.107 | -0.022, 0.893 | 0.031, 0.847 | 0.045, 0.781 | -0.323, 0.045 |
| **Gray Matter (Left)** | 0.391,<br>0.013 | -0.217, 0.178 | 0.137, 0.340 | 0.015, 0.927 | -0.323, 0.045 |
| **Cingulate Cortex (Left)** | 0.267,<br>0.095 | -0.177, 0.273 | 0.293, 0.067 | -0.090, 0.581 | -0.206, 0.208 |
| **Mean of All Regions** | 0.405,<br>0.010 | -0.217, 0.179 | 0.179, 0.269 | -0.069, 0.673 | -0.440, 0.005 |

**Table S2.** Spearman correlation coefficients and p-values between placebo-adjusted volume percent change and placebo-adjusted clinical score percent change at Week 39, reported for seven brain regions of interest and their cross-region mean (N = 40, except MoCA: N =39)

| **Table S2** |  |  |  |  |  |
| --- | --- | --- | --- | --- | --- |
|  | **ADAS-Cog13**<br>(Rho, p) | **ADCS-ADL**<br>(Rho, p) | **CDR-SB**<br>(Rho, p) | **MMSE**<br>(Rho, p) | **MoCA**<br>(Rho, p) |
| **Frontal Cortex (Left)** | -0.179, 0.257 | 0.370, 0.016 | -0.266, 0.089 | 0.346, 0.025 | 0.265, 0.094 |
| **Hippocampus (Left)** | -0.228, 0.147 | 0.236, 0.132 | -0.157, 0.321 | 0.255, 0.103 | 0.116, 0.472 |
| **Temporal Cortex (Left)** | -0.273, 0.080 | 0.464, 0.002 | -0.281, 0.071 | 0.471, 0.0016 | 0.403, 0.0089 |
| **Parietal Cortex (Left)** | -0.115, 0.467 | 0.196, 0.213 | -0.140, 0.377 | 0.256, 0.102 | 0.268, 0.090 |
| **Occipital Cortex (Left)** | -0.187, 0.236 | 0.297, 0.056 | -0.050, 0.755 | 0.494, 0.0009 | 0.417, 0.0066 |
| **Cingulate Cortex (Left)** | -0.014, 0.930 | 0.390, 0.011 | -0.176, 0.264 | 0.365, 0.017 | 0.136, 0.397 |
| **Mean of All Regions** | -0.196, 0.212 | 0.353, 0.022 | -0.162, 0.305 | 0.411, 0.0068 | 0.280, 0.076 |

